# Development and Validation of a Machine Learning Wrist-worn Step Detection Algorithm with Deployment in the UK Biobank

**DOI:** 10.1101/2023.02.20.23285750

**Authors:** Scott R. Small, Shing Chan, Rosemary Walmsley, Lennart von Fritsch, Aidan Acquah, Gert Mertes, Benjamin G. Feakins, Andrew Creagh, Adam Strange, Charles E. Matthews, David A. Clifton, Andrew J. Price, Sara Khalid, Derrick Bennett, Aiden Doherty

**Author notes:** **Correspondence to:** Aiden Doherty, PhD, Professor of Biomedical Informatics, University of Oxford, Nuffield Department of Population Health, Big Data Institute, Old Road Campus, Oxford, UK OX3 7LF.

## Abstract

**Background:** Step count is an intuitive measure of physical activity frequently quantified in a range of health-related studies; however, accurate quantification of step count can be difficult in the free-living environment, with step counting error routinely above 20% in both consumer and research-grade wrist-worn devices. This study aims to describe the development and validation of step count derived from a wrist-worn accelerometer and to assess its association with cardiovascular and all-cause mortality in a large prospective cohort study.

**Methods:** We developed and externally validated a hybrid step detection model that involves self-supervised machine learning, trained on a new ground truth annotated, free-living step count dataset (OxWalk, n=39, aged 19-81) and tested against other open-source step counting algorithms. This model was applied to ascertain daily step counts from raw wrist-worn accelerometer data of 75,493 UK Biobank participants without a prior history of cardiovascular disease (CVD) or cancer. Cox regression was used to obtain hazard ratios and 95% confidence intervals for the association of daily step count with fatal CVD and all-cause mortality after adjustment for potential confounders.

**Findings:** The novel step algorithm demonstrated a mean absolute percent error of 12.5% in free-living validation, detecting 98.7% of true steps and substantially outperforming other recent wrist-worn, open-source algorithms. Our data are indicative of an inverse dose-response association, where, for example, taking 6,596 to 8,474 steps per day was associated with a 39% [24-52%] and 27% [16-36%] lower risk of fatal CVD and all-cause mortality, respectively, compared to those taking fewer steps each day.

**Interpretation:** An accurate measure of step count was ascertained using a machine learning pipeline that demonstrates state-of-the-art accuracy in internal and external validation. The expected associations with CVD and all-cause mortality indicate excellent face validity. This algorithm can be used widely for other studies that have utilised wrist-worn accelerometers and an open-source pipeline is provided to facilitate implementation.

**Funding Acknowledgements:** This research has been conducted using the UK Biobank Resource under Application Number 59070. This research was funded in whole or in part by the Wellcome Trust [223100/Z/21/Z]. For the purpose of open access, the author has applied a CC-BY public copyright licence to any author accepted manuscript version arising from this submission. AD and SS are supported by the Wellcome Trust. AD and DM are supported by Swiss Re, while AS is an employee of Swiss Re. AD, SC, RW, SS, and SK are supported by HDR UK, an initiative funded by UK Research and Innovation, Department of Health and Social Care (England) and the devolved administrations. AD, DB, GM, and SC are supported by NovoNordisk. AD is supported by the BHF Centre of Research Excellence (grant number RE/18/3/34214). SS is supported by the University of Oxford Clarendon Fund. DB is further supported by the Medical Research Council (MRC) Population Health Research Unit. DC holds a personal academic fellowship from EPSRC. AA, AC and DC are supported by GlaxoSmithKline. SK is supported by Amgen and UCB BioPharma outside of the scope of this work. Computational aspects of this research were funded from the National Institute for Health Research (NIHR) Oxford Biomedical Research Centre (BRC) with additional support from Health Data Research (HDR) UK and the Wellcome Trust Core Award [grant number 203141/Z/16/Z]. The views expressed are those of the author(s) and not necessarily those of the NHS, the NIHR or the Department of Health.

## Introduction

Physical activity has been associated with lower risk of a wide range of non-communicable diseases and is a key feature of public health guidelines for cardiovascular health^1–3^. While researchers most commonly report device-measured activity in terms of overall acceleration or time-use behaviours derived from intensity thresholds^4^, the reporting of steps is a more intuitive measure of physical activity intrinsically linked to the key biomechanical feature of human gait^5^. However, current methods to measure steps from wrist-worn monitors during free-living activity are inaccurate^6^.

Most activity tracking devices with embedded step counting rely on proprietary step counting methods without transparent evaluation^7^, and many popular open-source step counting algorithms were not developed in accordance with, or lack validation against, direct observation ground truth step counts in a free-living environment^8–10^. Current standards require commercial activity trackers to estimate step counts with an error of less than 10% in laboratory-controlled treadmill testing^11^. Subsequently, many devices and algorithms perform well during scripted, moderately paced walking in controlled conditions^12,13^. However, step counting performance substantially deteriorates in the real-world environment, wherein mean absolute percent error (MAPE) is regularly well above 20% in both commercial and research-grade activity monitors during free living^6^. As a consequence, uncertainty exists around the strength and shape of the association of daily step count with all-cause mortality and cardiovascular mortality^14,15^, where recent studies have not used transparent or robustly validated free-living step counting algorithms.

In response, we set out to develop and validate a method to accurately measure steps in free-living environments. The purpose of this study was threefold: 1) to develop a novel self-supervised learning step detection algorithm trained with free-living stepping data, 2) to externally validate the algorithm alongside other open-source algorithms, and 3) to evaluate the face validity of this method in a large scale prospective cohort study by associating step counts with fatal CVD and all-cause mortality.

## Methods

### Development of the Free-Living, Ground Truth Annotated OxWalk Dataset

To develop the OxWalk^16^ dataset, participants contributed activity data during unscripted, free living. Participants wore four triaxial accelerometers (AX3, Axivity, Newcastle, UK), two placed side-by-side on the dominant wrist and two clipped to the dominant-side hip at the midsagittal plane. Accelerometers were synchronised using the Open Movement GUI software (v.1.0.0.42), with one recording at 100 Hz and the other at 25 Hz at each body location. Final accelerometer data was resampled to the nominal sampling rate and calibrated to local gravity using the Open Movement software package. Foot-facing video was captured using an action camera (Action Camera CT9500, Crosstour, Shenzhen, China) mounted at the participant’s beltline (Supplemental Figure 1). Participants were instructed to wear the camera for one hour and could remove the camera any time they felt uncomfortable or required additional privacy^17^. To create a clear, easily distinguishable data point for video and accelerometer synchronisation in this study, participants were asked to strike their accelerometers together with four forceful blows within camera view at the start of data collection^18^.

Ground truth annotation of steps was conducted within video annotation software (Elan 6.0, The Language Archive, Nijmegen, Netherlands) by two independent annotators (SS and LvF) blinded to each other’s results. Similar to Bassett et al., we identified the act of lifting a foot and placing it in a new location as a central tenant of step identification^5^. This definition was used as the framework for step annotation in the OxWalk dataset, with an annotated step being a repositioned foot linked to a change in gross body position along the floor. Annotated steps did not include foot shuffling, changing of foot alignment via pivoting, or shifting of weight from one foot to the other. Ethical approval for participant recruitment was obtained from the Central University Research Ethics Committee of the University of Oxford (Ref: R63137/RE001). Written informed consent was obtained from adult volunteers (aged 18 and above) with no lower limb injury within the previous six months and who were able to walk without an assistive device.

### Model Development and Evaluation

To develop the proposed step count model, a hybrid machine learning and peak detection algorithm was created wherein an activity classification model was first used to detect periods of walking and non-walking, followed by step counting only on predicted walking data epochs (Figure 1). Activity classification was performed using a self-supervised deep learning model developed by Yuan et al^20^ incorporating an 18-layer ResNet-V2^21^ pre-trained using self-supervised tasks on the UK Biobank accelerometer dataset. This pre-training step has previously demonstrated consistent performance improvement for downstream activity recognition tasks against Random Forest activity classification^20^. The pre-trained self-supervised learning model was then trained for supervised gait classification using the OxWalk dataset, wherein training data consisted of 10 second epochs of accelerometer data with ground-truth walk or non-walk labels. In the OxWalk dataset, walking was defined as at least four steps within the 10 second epoch. Ten-fold cross-validation was used to train and validate the walking activity classifier and evaluate end-to-end performance of the step detection pipeline. The participant dataset was divided into 10 equal random folds where one fold was left out for testing and the remaining folds underwent a randomised 80%-20% split for training and validation, respectively. Folds were stratified by class label and data was grouped by participant. The self-supervised learning model was trained on the remaining set with an early-stopping mechanism on the validation set when the loss stopped decreasing for 5 consecutive training epochs. The weights prior to early stopping were used to perform activity prediction on the test and validation set. An additional data-augmentation step was performed during training, whereby each triaxial training sample was randomly transformed with a rotation along a random axis and the axes were switched in a random order to make the model rotation invariant. The model was trained using PyTorch 1.12.1 and Adam optimisation^22^ with a learning rate of 0.0001. Weighted cross entropy loss was used, with the class weights set in such a way that the balance of walking and non-walking segments was 10% to 90%, respectively, bringing the class balance in line with 24-hour direct observation during free living in a previously collected dataset^23^. Finally, predictions on the validation set and corresponding ground-truth labels were used to train a Hidden Markov Model smoother which was then applied to the predictions in the test set.

**Figure 1:**
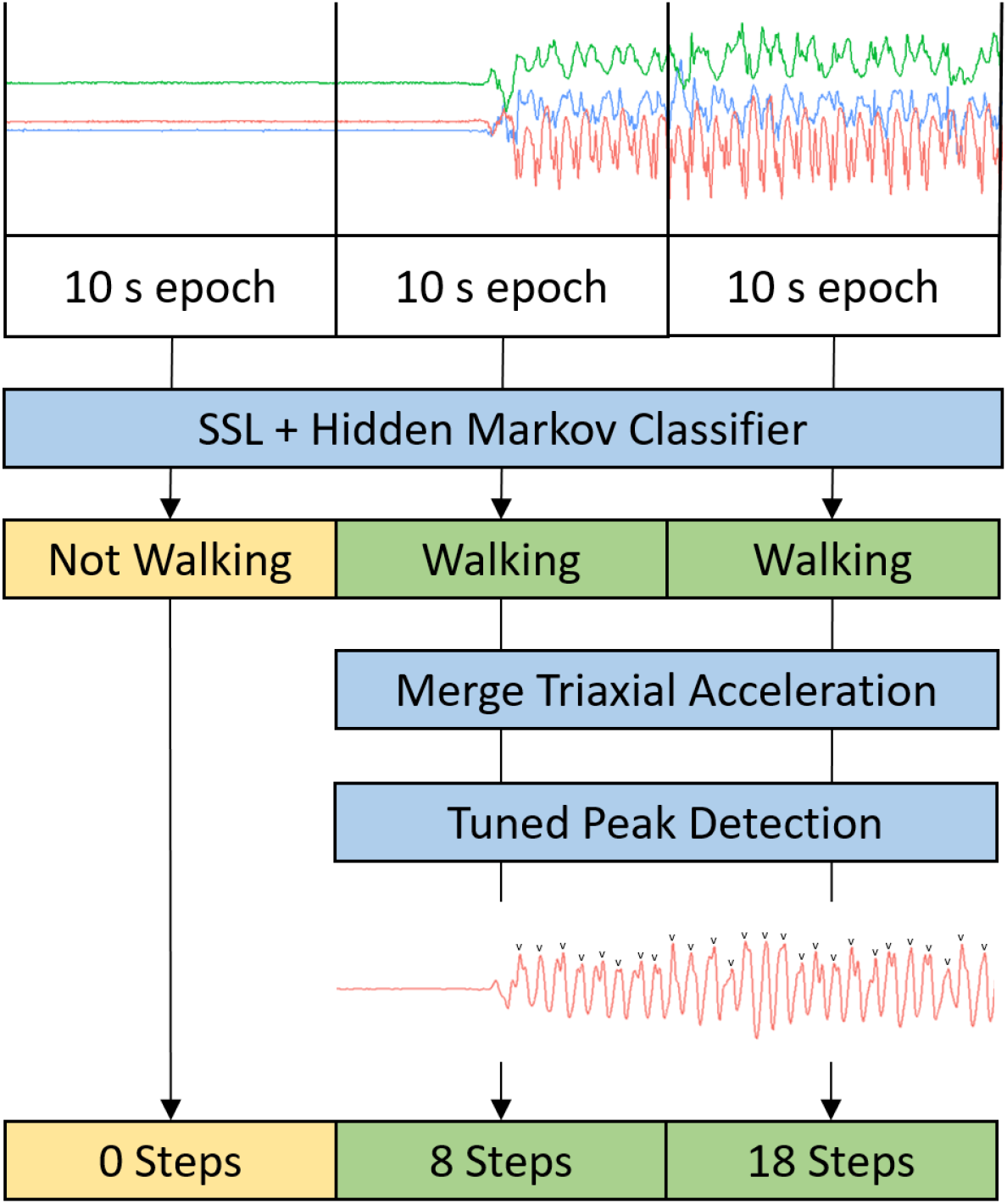
**Schematic of the process for generating step count from 30 seconds of raw triaxial accelerometer data using a hybrid self-supervised learning (SSL) and peak detection step counting model**.

Step counting was performed through peak detection on classified walking time windows using the “find_peaks” method from the SciPy Python package^24^. Euclidean norm of triaxial acceleration, minus 1 *g* to remove the effect of gravity, was clipped between ±2 *g* and lowpass filtered at 5 Hz prior to use as the input signal for peak detection. The “find_peaks” method detects local peaks using predefined heuristics including the minimum peak height (prominence), maximum peak width (width), and minimum time between peaks (distance). These heuristics served as detection hyperparameters for which optimal values would minimise the mean absolute error for step count in the validation set. Detection parameters were iterated across a pre-selected range of values (prominence: 0.1 to 1 *g*; distance: 0.2 to 2 s; width: 10 ms to 1 s).

Model performance metrics were calculated on participants within each test set; mean precision, recall, F1, Cohen’s kappa, and accuracy were used to evaluate walking classification, while MAPE and mean bias and Spearman’s rank correlation coefficient were calculated against ground truth step annotations. Following internal model validation, the final activity prediction model was retrained on the entire OxWalk dataset with an 80%-20% training-validation split prior to external deployment.

### External Model Validation

External model performance was assessed by applying the step detection algorithm to wrist-worn accelerometer data to an open-source, step-annotated dataset from Clemson University^19^. Within this external dataset, 30 participants contributed a mean of 37 minutes of activity, split between three distinct sessions of regular walking (two laps around a predefined path), semiregular walking (locating objects throughout a building), and irregular walking (collecting and assembling building blocks distributed around a room). Participants were video recorded throughout scripted activities, allowing timestamp-annotated steps while wearing Shimmer3 inertial measurement units (Shimmer, Dublin, Ireland) recording at 15 Hz. Researchers annotated steps as well as “shifts”, foot movement not necessarily tied to a change in body position, though these annotated shifts were not included in the current analysis^19^. Prediction error was quantified by calculating MAPE and mean percent under/overcounting bias for each gait subtype and overall, at the participant level, across all gait subtypes. Bland-Altman plots were created for comparison between cumulative ground truth and predicted step counts for each participant.

### Open-source Step Count Algorithm Assessment

In addition to assessment of the novel algorithm, two additional step counting approaches were evaluated in this study using both the OxWalk and Clemson datasets: 1) a recently-published acceleration-threshold algorithm by Ducharme et al. ^8^, and 2) the Verisense algorithm, a popular open-source peak detection algorithm developed from the Clemson dataset^25^ and previously applied to UK Biobank accelerometer data using integration with the GGIR package^26,27^. Further details for these algorithms are presented in Supplemental Note 1, while details of all datasets used are presented in Supplemental Table 1.

### Model Implementation into the UK Biobank

The UK Biobank is a prospectively recruited observational cohort of over 500,000 participants aged 40–69 at the time of recruitment, from 2006–2010^28^. From 2013–2015, participants were invited to wear an Axivity AX3 accelerometer on their dominant wrist, recording at 100 Hz, for a seven-day, 24 hours per day activity measurement window. In the current study, raw accelerometer data was processed from 103,391 available participants, after which data was excluded from participants with fewer than 72 hours of wear, those lacking data across the entire diurnal cycle, with poor device calibration, or with unrealistic average acceleration (>100 m*g*)^4^.

The externally validated hybrid SSL step detection model was applied to raw accelerometer data from the UK Biobank. Overall daily step count was reported as the median number of steps taken across the seven-day measurement period. Missing step count data from non-wear was imputed by averaging step count from the corresponding time of day in all other valid days, similar to the imputation of vector magnitude acceleration during non-wear in the UK Biobank physical activity cohort^4^. One-minute peak cadence was calculated as previously described by Saint-Maurice et al^29^.

### Statistical Analysis

UK Biobank participants with prevalent cardiovascular disease or cancer as a primary diagnosis, as identified by International Classification of Diseases (ICD) codes I00–I99 and C00–C97 in their routine hospital data, were removed from analysis. Spearman’s rank correlation (r) was calculated between step count, peak cadence, overall acceleration, and UK Biobank derived activity time use activity classification^23^. Daily step count and one minute peak cadence were stratified across demographic and self-reported health variables as collected by the UK Biobank at the time of enrolment. Analysis of variance and Tukey Honestly Significant Difference tests were conducted to compare step count based on self-reported health and usual walking pace.

Multivariable adjusted estimates of the effect of quintiles of step count on the relative hazards of cardiovascular mortality and all-cause mortality were derived using Cox proportional hazards regression using age as the underlying timescale^30,31^. Date and cause of death was gathered from the UK Biobank linked death registry. Length of follow-up was calculated from censoring dates from the data sources or date of death. Further detail is provided in Supplemental Notes 2-3. Step count detection was deployed on the UK Biobank using the University of Oxford Biomedical Research Computing cluster, while statistical analysis was completed using R (v.4.1.1) on the UK Biobank Research Analysis Platform. Statistical code is available at https://github.com/OxWearables/UKB_steps_mortality.

## Results

### Step Count Validation in the OxWalk Dataset

Accelerometer and ground truth camera data was collected from 39 participants (19 female, 20 male) with a mean age of 38.5 years (range 19.5 to 81.2 years), a mean wear time of 58 minutes, and a median [interquartile range (IQR)] 863 [312–2,123] steps within the measurement period. Thirty-three participants were annotated by both annotators, resulting in a corresponding step count MAPE of 4.0% and interclass correlation coefficient of 1.0 between annotators. Internal validation of the self-supervised learning model identified bouts of walking with a Cohen’s Kappa performance of 0.79 (Supplemental Table 3). Overall cross-validation of step detection in the self-supervised learning model resulted in a 12.5% MAPE, 1.3% underestimation of steps, and correlation of r = 0.98 against ground truth in the free-living OxWalk dataset. For comparison, external validation of the step counting of the 100 Hz OxWalk wrist-worn dataset using the Ducharme acceleration-threshold algorithm^8^ resulted in a 69.1% overestimation of steps (231.3 % MAPE, r= 0.91) across all participants. External validation of the Verisense algorithm^10,25^, incorporated into recent UK Biobank papers^14,26^, produced a 63.5% MAPE, 7.2% underestimation bias, and r = 0.85 against free-living ground truth step counts (Supplemental Table 2). Bland-Altman plots for model comparisons against ground truth OxWalk step count are presented in Figure 2, demonstrating lower variability and tighter agreement with ground truth using the novel step detection algorithm in the free-living dataset.

**Figure 2:**
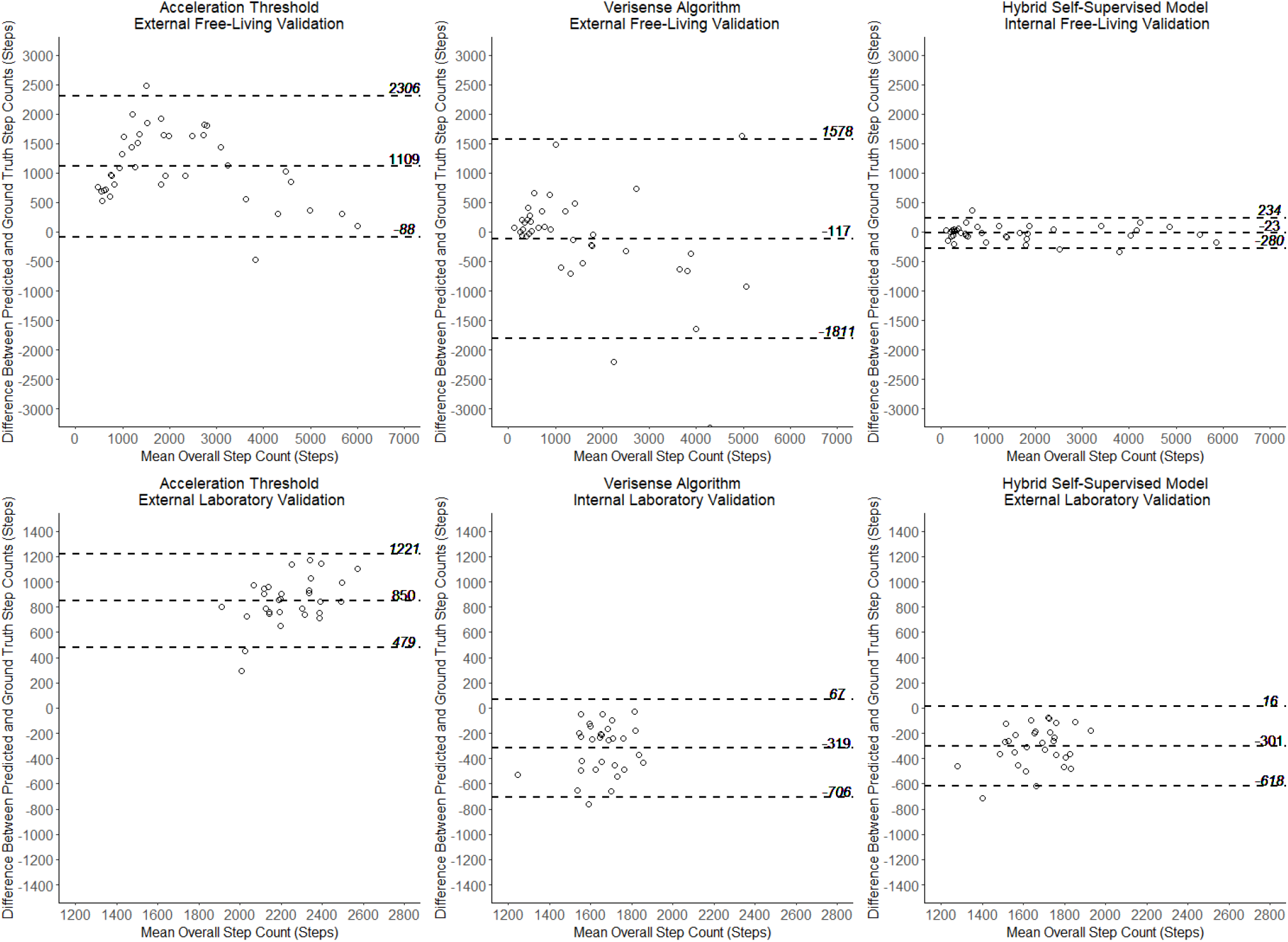
**Bland-Altman plots with dotted 95% limits of agreement for the comparison of step counting models in the (Top) OxWalk free-living dataset of 39 adults and (Bottom) Clemson laboratory-based dataset of 30 young adults. Left: baseline acceleration threshold model**^8^, **Centre: Verisense algorithm**^25^, **and Right: the novel hybrid self-supervised learning model**.

### Step Count Validation in the Clemson Dataset

Bland-Altman plots for the performance of each prediction method in the overall Clemson dataset are also presented in Figure 2. This plot again demonstrates reduced variability and bias against ground truth using the novel model compared to reference algorithms. In external validation, the threshold model by Ducharme et al.^8^ performed well during sessions of regular gait, but poorly irregular gait, culminating in an overall MAPE of 47.5% and a 46.9% overestimation of steps at the participant-level, across all gait subtypes. The Verisense algorithm, for which this dataset serves as an internal validation, demonstrated a 17.6% underestimation of steps and a 17.3% per-participant MAPE over all gait subtypes, including 16.3% MAPE during regular walking (Supplemental Table 4). External validation of our novel self-supervised learning hybrid step algorithm performed best in the Clemson dataset, producing a 16.5% MAPE and 16.6% underestimation across all gait subtypes, including 9.2% MAPE during regular walking. Due to superior performance in free-living and laboratory-based validation, the SSL step detection model was selected for analysis of UK Biobank data.

### Step Counts in the UK Biobank Physical Activity Cohort

Baseline data from 75,493 UK Biobank participants without prevalent CVD or cancer is presented in Table 1 and Supplemental Figure 2. Peak step cadence demonstrated expected variations by self-reported usual walking pace (Supplemental Figure 3) and our measurements of steps demonstrated orthogonality to standard overall acceleration and time-use metrics (Supplemental Figure 4). Participants that self-reported that their overall health was excellent were more active than all other participants, taking 2,947 more steps [95% CI 2,678–3,215] (p < 0.001) than those reporting that their overall health was poor. Similarly, self-reported brisk walkers had a peak one-minute cadence 11.2 steps per minute [95% CI 10.6–11.7] (p < 0.001) higher than slow walkers. Adjusted mean daily step counts by self-reported health status and by selected physician-diagnosed chronic conditions are presented in Figure 3.

**Table 1:** Overall Physical Activity Metrics by Demographic Characteristic in the UK Biobank.

| Characteristic | N (%) | Daily Steps | Peak Cadence<br>(Steps per<br>minute) | Overall Acceleration (mg) |
| --- | --- | --- | --- | --- |
| Overall | 75,493 (100.0) | 9,352 [7,099-11,973] | 117 [111-122] | 27.5 [22.9-32.9] |
| Sex |  |  |  |  |
| Female | 43,802 (58.0) | 9,267 [7,050-11,840] | 118 [113-124] | 27.8 [23.3-33.2] |
| Male | 31,691 (42.0) | 9,468 [7,182-12,159] | 114 [109-119] | 27.0 [22.3-32.6] |
| Age, years |  |  |  |  |
| 40-49 | 7,229 (9.6) | 9,412 [7,181-12,047] | 119 [113-125] | 30.4 [25.5-36.5] |
| 50-59 | 23,390 (31.0) | 9,348 [7,136-12,014] | 118 [113-124] | 29.1 [24.4-34.8] |
| 60-69 | 32,903 (43.6) | 9,497 [7,222-12,122] | 116 [110-122] | 26.9 [22.5-32.1] |
| 70-79 | 11,971 (15.9) | 8,925 [6,667-11,374] | 114 [108-120] | 24.6 [20.5-29.2] |
| Ethnicity |  |  |  |  |
| Nonwhite | 2,380 (3.2) | 9,050 [6,764-11,691] | 118 [111-124] | 28.7 [23.9-34.2] |
| White | 73,113 (96.8) | 9,362 [7,115-11,981] | 116 [111-122] | 27.4 [22.9-32.9] |
| Body Mass Index |  |  |  |  |
| Underweight (<18.5<br>kg/m<sup>2</sup>) | 444 (0.6) | 10,000 [7,972-13,120] | 121 [115-127] | 31.3 [25.2-36.8] |
| Normal weight (18.5-24.9<br>kg/m<sup>2</sup>) | 30,312 (40.2) | 9,923 [7,696-12,526] | 119 [113-125] | 29.5 [24.7-35.1] |
| Overweight (25.0-29.9<br>kg/m<sup>2</sup>) | 30,791 (40.8) | 9,363 [7,174-11,943] | 116 [110-121] | 27.0 [22.7-32.1] |
| Obese (30+ kg/m<sup>2</sup>) | 13,946 (18.5) | 7,956 [5,896-10,457] | 113 [107-119] | 24.4 [20.3-29.2] |
| Education |  |  |  |  |
| School Leaver | 16,710 (22.1) | 8,933 [6,749-11,532] | 116 [110-122] | 27.0 [22.3-32.5] |
| Further Education | 25,052 (33.2) | 9,172 [6,908-11,846] | 116 [110-122] | 27.5 [22.9-32.9] |
| Higher Education | 33,731 (44.7) | 9,679 [7,454-12,248] | 117 [112-123] | 27.7 [23.2-33.1] |
| Smoking Status |  |  |  |  |
| Never | 44,231 (58.6) | 9,422 [7,200-12,007] | 117 [112-123] | 27.8 [23.2-33.2] |
| Former | 26,107 (34.6) | 9,329 [7,043-11,984] | 116 [110-122] | 27.3 [22.7-32.7] |
| Current | 5,155 (6.8) | 8,784 [6,532-11,547] | 114 [109-120] | 26.3 [21.5-31.8] |
| Alcohol Consumption |  |  |  |  |
| Never | 4,086 (5.4) | 8,906 [6,448-11,620] | 116 [109-122] | 26.8 [21.8-32.4] |
| < 3 Days Per Week | 34,319 (45.5) | 9,031 [6,810-11,622] | 117 [111-122] | 27.3 [22.6-32.7] |
| 3+ Days Per Week | 37,088 (49.1) | 9,685 [7,475-12,281] | 117 [111-122] | 27.8 [23.3-33.2] |
| Townsend Deprivation |  |  |  |  |
| Least Deprived (<-3.8) | 18,854 (25.0) | 9,332 [7,200-11,945] | 116 [110-122] | 27.6 [23.1-32.9] |
| Second Least Deprived (-<br>3.8 to -2.2) | 18,892 (25.0) | 9,326 [7,145-11,860] | 116 [111-122] | 27.5 [23.0-32.9] |
| Second Most Deprived (-<br>2.5 to -1.2) | 18,869 (25.0) | 9,362 [7,080-11,951] | 117 [111-122] | 27.5 [22.9-32.9] |
| Most Deprived (≥-0.2) | 18,878 (25.0) | 9,384 [6,974-12,124] | 117 [111-124] | 27.4 [22.6-32.9] |
| Self-Reported Usual Walking Pace |  |  |  |  |
| Brisk | 36,733 (48.7) | 9,787 [7,567-12,405] | 118 [113-124] | 29.0 [24.4-34.6] |
| Steady | 35,727 (47.3) | 9,078 [6,890-11,653] | 115 [110-121] | 26.4 [22.0-31.4] |
| Slow | 2,905 (3.8) | 6,889 [4,582-9,495] | 109 [102-116] | 22.3 [18.1-27.2] |
| None of the above | 61 (0.1) | 5,773 [3,371-10,301] | 107 [99-112] | 22.5 [18.4-28.4] |
| Missing | 67 (0.1) | 3,148 [868-5,758] | 96 [65-105] | 18.9 [14.1-24.2] |
| Self-Reported Overall Health |  |  |  |  |
| Excellent | 17,781 (23.6) | 9,855 [7,708-12,432] | 118 [113-124] | 29.2 [24.5-35.0] |
| Good | 45,755 (60.6) | 9,397 [7,178-12,017] | 116 [111-122] | 27.5 [23.0-32.7] |
| Fair | 10,520 (13.9) | 8,468 [6,208-11,099] | 114 [108-120] | 25.3 [20.9-30.4] |
| Poor | 1,437 (1.9) | 6,939 [4,394-9,626] | 110 [103-117] | 22.9 [18.4-28.1] |
| Wear Season |  |  |  |  |
| Spring | 17,327 (23.0) | 9,479 [7,193-12,130] | 117 [111-123] | 27.9 [23.2-33.4] |
| Summer | 20,014 (26.5) | 9,812 [7,525-12,520] | 116 [110-121] | 28.1 [23.4-33.6] |
| Autumn | 22,342 (29.6) | 9,236 [7,040-11,804] | 117 [111-123] | 27.4 [22.9-32.8] |
| Winter | 15,810 (20.9) | 8,774 [6,647-11,288] | 117 [111-123] | 26.6 [22.1-31.7] |

*Activity metrics reported as unadjusted median [interquartile range]*

**Figure 3:**
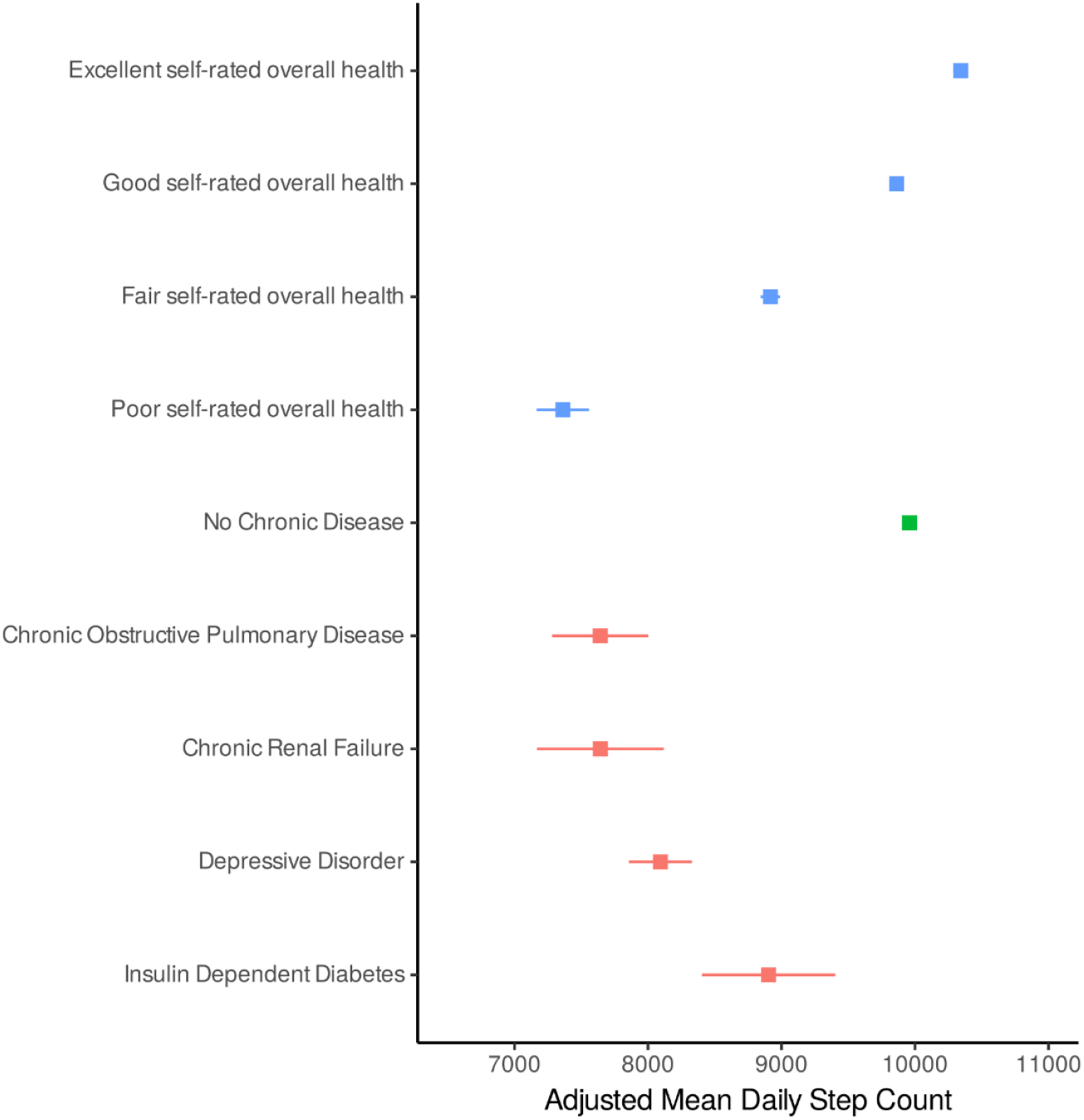
**Adjusted estimated marginal mean (95% confidence interval) daily step count according to self-reported overall health status, hospital data derived chronic disease status, and select diagnoses for 75**,**493 UK Biobank participants**. Mean daily step counts are adjusted for age and sex.

### Association of Step Counts with All-Cause and Cardiovascular Mortality

The Cox regression analysis cohort had a median follow-up of 6.9 [IQR 6.3–7.4] years, with 572 events in the CVD mortality analysis and 1,844 events in the all-cause mortality analysis (Figure 4). For CVD mortality, a curvilinear association was observed with a linear association observed between the first and third fifths of the step count distribution and then a flattening of the association for the top two fifths of the distribution. For example, a median daily step count of 8,474 to 10,284 steps per day was associated with a 56% [43–66%] lower risk of CVD mortality compared to participants taking fewer than 6,596 steps per day, whereas taking 12,677 or more steps was associated with a 56% [43-66%] lower risk on CVD mortality. Similar results were observed in the analysis of all-cause mortality and median daily step count, with a 39% [30-47%] and 43% [34-51%] lower risk of all-cause mortality in the middle and most active 20%, respectively.

**Figure 4:**
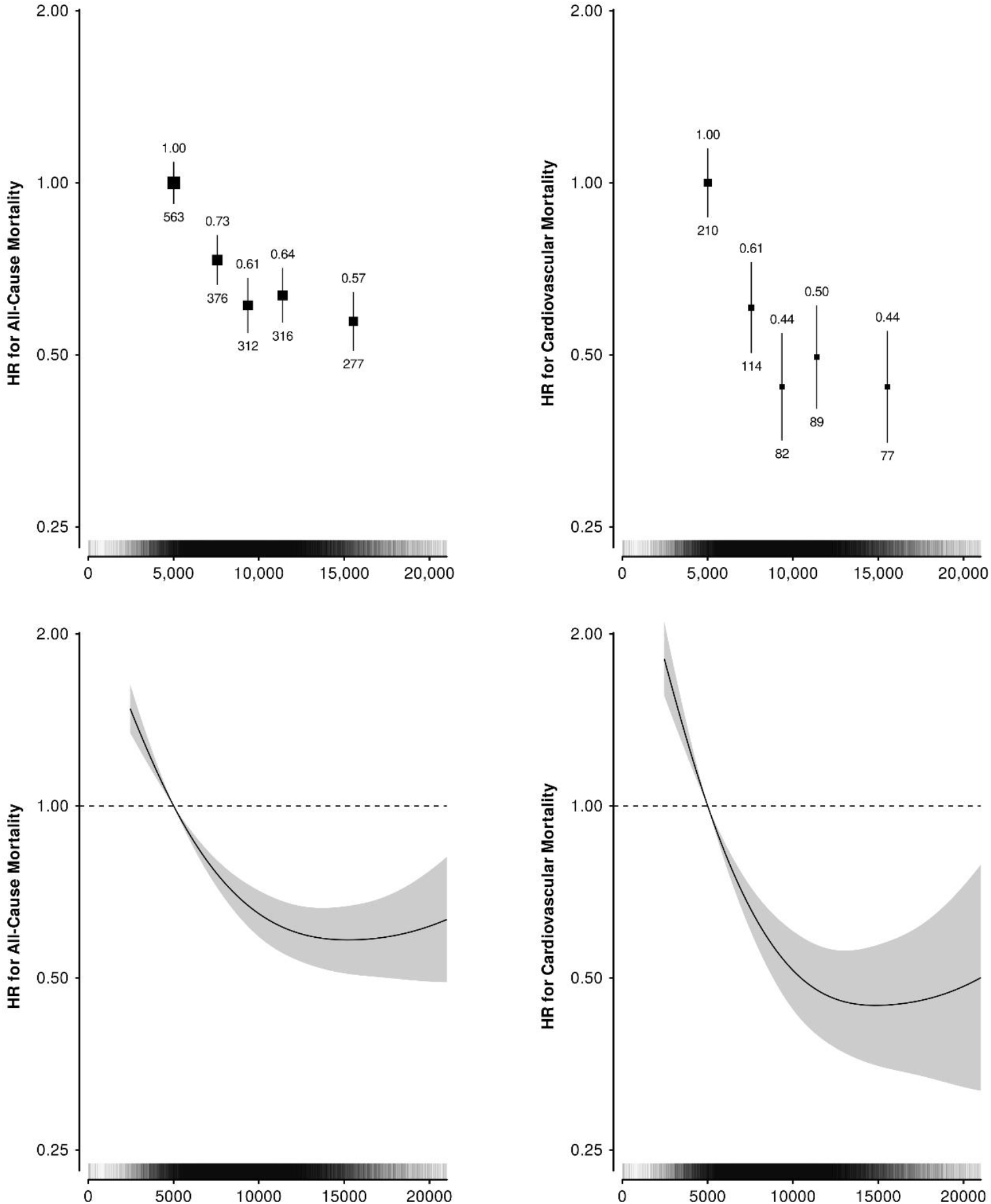
**(Top) Forest plots for all-cause mortality and cardiovascular disease mortality associations with quintiles of daily step count, (Bottom) continuous daily step count for 75**,**493 UK Biobank participants**. Hazard ratios (HR) and 95% confidence intervals were calculated using age as a timescale, adjusted for sex, ethnicity, education, alcohol intake, smoking status, Townsend deprivation index, processed meat intake, fresh fruit intake, oily fish intake, and added salt intake. HR is above and number of events is plotted below each data point. Spline plot of hazard ratio and 95% confidence interval of the association of continuously modelled median daily step count. Vertical bars along the step axis indicate distribution of participant daily step counts.

## Discussion

We have developed a new open-source step counting method, informed by self-supervised machine learning methods that substantially outperforms current wrist-worn step counting algorithms in the free-living environment. The open data and code released with this manuscript will provide the global research community access to a more transparent and well-validated method to measure steps in large-scale wrist-worn accelerometer datasets. When applying the algorithm and resulting step metric in epidemiological analysis, we demonstrated that a higher daily step count is associated with a lower risk of all-cause and cardiovascular mortality.

Our novel approach of using a hybrid step detection model that involves self-supervised machine learning outperformed existing wrist-worn step counting methods, producing a 12.5% MAPE and 1.3% step underestimation during free living. Wrist-worn step counting is highly popular in both commercial and research applications, but valid step detection at the wrist can be associated with high measurement error relative to ground truth. In 2018, Toth et al.^6^ assessed wrist-worn step detection in free-living conditions, finding error rates between 18% and 120% across a range of methodologies. We found similar performance in current open-source algorithms during free-living testing, with a mean average percent error ranging from 64% to 231%. Even while analysing data from a different device and sampling rate, external validation of the novel model in the Clemson dataset demonstrated a 9.2% error during regular walking in the laboratory-based setting, below the 10% MAPE threshold required during treadmill-based validation^11^. External validation of the novel model outperformed both reference algorithms, including the Verisense algorithm, which was trained and tuned using the Clemson laboratory dataset^10,25^.

This study demonstrates a strong inverse curvilinear association between increased step count and lower risk of fatal CVD and all-cause mortality while highlighting the importance of accurate step detection algorithms in epidemiological analysis. Our current results parallel those of Paluch et al.^15^, who demonstrated higher daily step counts are associated with an incrementally lower risk of all-cause mortality across 15 international longitudinal cohorts nearly exclusively using hip-mounted devices. Using less accurate step-detection methods, another study has also indicated a curvilinear association between daily steps and CVD mortality^27^. Though the direction of epidemiological associations may remain broadly similar across step detection algorithms, it is important that algorithms derive step counts as accurately as possible. Accurate step counting will be particularly important when translating results into target levels of physical activity in guidelines compatible with device-measured activity^32^. Reporting of inaccurate step counts may additionally be demotivating and counterproductive in terms of health metrics and behavioural change for individuals monitoring their own physical activity^33^.

Clear strengths of our study include the development of a step counting algorithm trained in a large dataset of free-living, wrist-worn accelerometer data with doubly-annotated ground truth video and demonstrated high accuracy. While this training data consisted of short 1-hour data collection windows, it is important to note that the current study algorithm is trained on one of the most complete free-living, open-source datasets to date. Some overestimation of step counts may occur when applied to multiday protocols due to the lack of extended periods of sedentary inactivity in the short training data, however; class rebalancing was utilised to minimise this effect. In the future, it will be important to further assess the robustness of this method across a variety of populations and against 24-hour free-living, ground truth annotated step count data.

## Conclusions

We have developed a new, open, and transparent method that markedly improves the ability to measure steps in large-scale wrist-worn accelerometer datasets. While using this validated step detection method trained using free-living data, we demonstrate an inverse dose response of daily step count with all-cause and cardiovascular disease mortality. This reinforces public health messaging of “the more, the better” approaches toward step count guidelines, encouraging any increase in physical activity, particularly in populations wherein a specific target number of daily steps may be unrealistic or feel unreachable.

## Data Availability

The OxWalk dataset generated in this study is available for download and free for use through the Oxford University Research Archive (https://ora.ox.ac.uk/objects/uuid:19d3cb34-e2b3-4177-91b6-1bad0e0163e7). An open-source accelerometer processing tool integrating the hybrid machine learning step detection method derived in this study is available for use at https://github.com/OxWearables/stepcount.

https://github.com/OxWearables/stepcount

https://github.com/OxWearables/UKB_steps_mortality

## Data Availability

The OxWalk dataset generated in this study is available for download and free for use through the Oxford University Research Archive (https://ora.ox.ac.uk/objects/uuid:19d3cb34-e2b3-4177-91b6-1bad0e0163e7). An open-source accelerometer processing tool integrating the hybrid machine learning step detection method derived in this study will be available for use at https://github.com/OxWearables/stepcount.

## References

1 World Health Organization. 2020 WHO guidelines on physical activity and sedentary behavior. Geneva: World Health Organization, 2020.

2 Khurshid S, Weng L-C, Nauffal V, et al. Wearable accelerometer-derived physical activity and incident disease. DOI:10.1038/s41746-022-00676-9.

3 Barker J, Smith Byrne K, Doherty A, et al. Physical activity of UK adults with chronic disease: cross-sectional analysis of accelerometer-measured physical activity in 96 706 UK Biobank participants. Int J Epidemiol 2019; published online Feb 5. DOI:10.1093/ije/dyy294.

4 Doherty A, Jackson D, Hammerla N, et al. Large scale population assessment of physical activity using wrist worn accelerometers: The UK biobank study. PLoS One 2017; 12: 1– 14.

5 Bassett DR, Toth LP, LaMunion SR, Crouter SE. Step Counting: A Review of Measurement Considerations and Health-Related Applications. Sport. Med. 2017; 47: 1303–15.

6 Toth LP, Park S, Springer CM, Feyerabend MD, Steeves JA, Bassett DR. Video-Recorded Validation of Wearable Step Counters under Free-living Conditions. Med Sci Sports Exerc 2018; 50: 1315–22.

7 Johnston W, Judice PB, García PM, et al. Recommendations for determining the validity of consumer wearable and smartphone step count: Expert statement and checklist of the INTERLIVE network. Br J Sports Med 2020; 0: 1–14.

8 Ducharme SW, Lim J, Busa MA, et al. A Transparent Method for Step Detection Using an Acceleration Threshold. J Meas Phys Behav 2021; 1: 1–10.

9 Femiano R, Werner C, Wilhelm M, Eser P. Validation of open-source step-counting algorithms for wrist-worn tri-axial accelerometers in cardiovascular patients. Gait Posture 2022; 92: 206–11.

10 Maylor BD, Edwardson CL, Dempsey PC, et al. Stepping towards More Intuitive Physical Activity Metrics with Wrist-Worn Accelerometry: Validity of an Open-Source Step-Count Algorithm. Sensors 2022, Vol 22, Page 9984 2022; 22: 9984.

11 ANSI/CTA-2056. Physical Activity Monitoring for Fitness Wearables: Step Counting. Consumer Technology Association, 2016 https://webstore.ansi.org/standards/ansi/cta20562016ansi.

12 Feito Y, Bassett DR, Thompson DL, Y F, DR B, DL T. Evaluation of activity monitors in controlled and free-living environments. Med Sci Sports Exerc 2012; 44: 733–41.

13 Mora-Gonzalez J, Gould ZR, Moore CC, et al. A catalog of validity indices for step counting wearable technologies during treadmill walking: the CADENCE-adults study. Int J Behav Nutr Phys Act 2022 191 2022; 19: 1–16.

14 Del Pozo Cruz B, Ahmadi MN, Lee IM, Stamatakis E. Prospective Associations of Daily Step Counts and Intensity with Cancer and Cardiovascular Disease Incidence and Mortality and All-Cause Mortality. JAMA Intern Med 2022; : 1DUMMY.

15 Paluch AE, Bajpai S, Bassett DR, et al. Daily steps and all-cause mortality: a meta-analysis of 15 international cohorts. Lancet Public Heal 2022; 7: e219–28.

16 Small SR, von Fritsch L, Doherty A, Khalid S, Price AJ. OxWalk: Wrist and hip-based activity tracker dataset for free-living step detection and gait recognition. 2022. DOI:10.5287/bodleian:ORQ2abnbR.

17 Kelly P, Marshall SJ, Badland H, et al. An ethical framework for automated, wearable cameras in health behavior research. Am J Prev Med 2013; 44: 314–9.

18 Fortune E, Lugade V, Morrow M, Kaufman K. Validity of using tri-axial accelerometers to measure human movement - Part II: Step counts at a wide range of gait velocities. Med Eng Phys 2014; 36: 659–69.

19 Mattfeld R, Jesch E, Hoover A. Evaluating pedometer algorithms on semi-regular and unstructured gaits. Sensors 2021; 21: 13–6.

20 Yuan H, Chan S, Creagh AP, Tong C, Clifton DA, Doherty A. Self-supervised Learning for Human Activity Recognition Using 700,000 Person-days of Wearable Data. arXiv 2022; published online June 6. DOI:10.48550/arxiv.2206.02909.

21 He K, Zhang X, Ren S, Sun J. Identity mappings in deep residual networks. Lect Notes Comput Sci (including Subser Lect Notes Artif Intell Lect Notes Bioinformatics) 2016; 9908 LNCS: 630–45.

22 Kingma DP, Ba JL. Adam: A method for stochastic optimization. In: 3rd International Conference on Learning Representations, ICLR 2015 - Conference Track Proceedings. International Conference on Learning Representations, ICLR, 2015. https://arxiv.org/abs/1412.6980 (accessed Dec 21, 2022).

23 Walmsley R, Chan S, Smith-Byrne K, et al. Reallocation of time between device-measured movement behaviours and risk of incident cardiovascular disease. BMJ Publishing Group Ltd and British Association of Sport and Exercise Medicine, 2021.

24 Virtanen P, Gommers R, Oliphant TE, et al. SciPy 1.0: fundamental algorithms for scientific computing in Python. Nat Methods 2020 173 2020; 17: 261–72.

25 Patterson MR. Verisense-Toolbox/Verisense_step_algorithm at master. ShimmerEngineering/Verisense-Toolbox. https://github.com/ShimmerEngineering/Verisense-Toolbox/tree/master/Verisense_step_algorithm (accessed Oct 20, 2022).

26 Del Pozo Cruz B, Ahmadi M, Naismith SL, Stamatakis E. Association of Daily Step Count and Intensity with Incident Dementia in 78430 Adults Living in the UK. JAMA Neurol 2022. DOI:10.1001/jamaneurol.2022.2672.

27 Del Pozo Cruz b, Ahmadi Mn, Lee ; I-Min, Stamatakis E. Prospective Associations of Daily Step Counts and Intensity With Cancer and Cardiovascular Disease Incidence and Mortality and All-Cause Mortality Supplemental content. JAMA Intern Med 2022; published online Sept 12. DOI:10.1001/JAMAINTERNMED.2022.4000.

28 Sudlow C, Gallacher J, Allen N, et al. UK Biobank: An Open Access Resource for Identifying the Causes of a Wide Range of Complex Diseases of Middle and Old Age. PLOS Med 2015; 12: e1001779.

29 Saint-Maurice PF, Troiano RP, Bassett DR, et al. Association of Daily Step Count and Step Intensity with Mortality among US Adults. JAMA - J Am Med Assoc 2020; 323: 1151–60.

30 Cox DR. Regression Models and Life-Tables. J R Stat Soc Ser B 2016; 15: 1–23.

31 Cologne J, Hsu WL, Abbott RD, et al. Proportional hazards regression in epidemiologic follow-up studies: An intuitive consideration of primary time scale. Epidemiology 2012; 23: 565–73.

32 Thompson D, Batterham AM, Peacock OJ, Western MJ, Booso R. Feedback from physical activity monitors is not compatible with current recommendations: A recalibration study. Prev Med (Baltim) 2016; 91: 389–94.

33 Zahrt OH, Evans K, Murnane E, et al. Effects of Wearable Fitness Trackers and Activity Adequacy Mindsets on Affect, Behavior, and Health: Longitudinal Randomized Controlled Trial. J Med Internet Res 2023;25e40529 https://www.jmir.org/2023/1/e40529 2023; 25: pe40529.

34 Gu F, Khoshelham K, Shang J, Yu F, Wei Z. Robust and accurate smartphone-based step counting for indoor localization. IEEE Sens J 2017; 17: 3453–60.

35 Lenth R V. emmeans: Estimated Marginal Means, aka Least-Squares Means. 2022. https://cran.r-project.org/package=emmeans.

36 Terry M. Therneau, Patricia M. Grambsch. Modeling Survival Data: Extending the Cox Model. New York: Springer, 2000.

37 Easton D, Peto J, AG B. Floating absolute risk: an alternative to relative risk in survival and case-control analysis avoiding an arbitrary reference group. Stat Med 1991; 10: 1025–35.

38 Carstensen B, Plummer M, Laara E, Hills M. Epi: A Package for Statistical Analysis in Epidemiology. 2022. https://cran.r-project.org/package=Epi.

