## Supplemental Materials for "Development and Validation of a Machine Learning Wrist-worn Step Detection Algorithm with Deployment in the UK Biobank"

Supplemental Table 1: Summary of Datasets ........................................................................ ii

Supplemental Table 2: Step Count Results and Performance Metrics Based in the OxWalk Dataset iii

Supplemental Table 3: Internal Validation Gait Classification Metrics for the Self-Supervised Learning Model ........................................................................ iv

Supplemental Table 4: Step Count Results and Performance Metrics Based in the External Clemson Shimmer3 Dataset Across all Participants and Gait Subtypes ........................................................................ v

Supplemental Figure 1: A foot-facing action camera clipped at the participant's waistline recording video for step count annotation. The participant is also wearing Axivity AX3 accelerometers at the dominant wrist and at the hip. ........................................................................ vi

Supplemental Figure 2: Flow diagram for descriptive and survival analyses in UK Biobank physical activity cohort. ........................................................................ vii

Supplemental Figure 3: Peak step cadence in the UK Biobank Physical Activity Cohort, summarised by self-reported walking pace and sex. ........................................................................ viii

Supplemental Figure 4: Spearman's rank correlation plot between accelerometer-based physical activity metrics, including overall acceleration, daily step count, and machine learning activity classifications of time spent in sleep, sedentary behaviour, light activity, and moderate-to-vigorous activity (MVPA). ........................................................................ ix

Supplemental Note 1: Comparative Step Detection Models ........................................................................ x

Supplemental Note 2: Statistical Analysis, Further Detail ........................................................................ xi

Supplemental Note 3: Field and Code Usage ........................................................................ xii

### Supplemental Table 1: Summary of Datasets

| Characteristic | OxWalk[16] | Clemson Shimmer3[19] | UK Biobank Physical Activity Cohort[4] |
|---|---|---|---|
| Institution | University of Oxford | Clemson University | UK Biobank |
| Number of Participants | 39 | 30 | ~100,000 |
| Sensor | Axivity AX3<br>triaxial accelerometer | Shimmer3<br>9-axis inertial measurement unit | Axivity AX3<br>triaxial accelerometer |
| Sampling Rate | 25 Hz and 100 Hz | 15 Hz | 100 Hz |
| Body Location | Dominant wrist and dominant hip | Non-dominant wrist, hip, and ankle | Dominant wrist |
| Activity Protocol | Unscripted<br>free-living | Laboratory scripted sessions of regular, semiregular, and irregular gait | Unscripted<br>free-living |
| Measurement Window | 1 hour | 37 minutes | 7 days |
| Ground Truth Capture | Foot-facing<br>video camera<br>worn at waist | Participant-facing video camera held<br>by researcher | None |
| Annotation of data | Timestamped steps by two annotators | Timestamped steps<br>and shifts | N/A |
| Label granularity | Timestamps to<br>0.001 s precision | Timestamps to<br>0.001 s precision | N/A |
| Participant Age | 38.5 (SD 14.0) years<br>Range: 19 - 81 | 21.9 (SD 2.4) years | 62.4 (SD 7.8) years<br>Range 43 - 79 |
| Participant Sex | 19 Female,<br>20 Male | 15 Female,<br>15 Male | 56% Female, 44% Male |

**Supplemental Table 2: Step Count Results and Performance Metrics Based in the OxWalk Dataset**

| | Ground Truth | Acceleration Threshold Model[8] | Verisense Algorithm[25] | SSL Hybrid Model |
|---|---|---|---|---|
| Total Steps | 62,782 | 106,160 | 58,356 | 62,025 |
| Mean Steps | 1,613 | 2,722 | 1,496 | 1,590 |
| Mean Bias (%) | | 69.1% | -7.2% | -1.3% |
| MAPE (SD) | | 231.3% (254.0%) | 63.5% (103.6%) | 12.5% (15.5%) |

*MAPE (SD): Mean Absolute percent error (standard deviation)*

**Supplemental Table 3: Internal Validation Gait Classification Metrics for the Self-Supervised Learning Model**

| | F1 | Cohen’s Kappa | Precision | Recall | Accuracy |
|---|---|---|---|---|---|
| SSL Hybrid Model | 0.842 ± 0.148 | 0.7890 ± 0.149 | 0.908 ± 0.088 | 0.807 ± 0.192 | 0.948 ± 0.047 |

**Supplemental Table 4: Step Count Results and Per-Participant Performance Metrics Based in the External Clemson Shimmer3 Dataset**

| **Overall Per-Participant** | Ground Truth | Acceleration Threshold[8] | Verisense[25] | SSL Hybrid Model |
|---|---|---|---|---|
| Total Steps | 54,387 | 79,885 | 44,813 | 45,357 |
| Mean Participant Steps | 1,813 | 2,663 | 1,494 | 1,512 |
| % Mean Bias | | +46.9% | -17.6% | -16.6% |
| MAPE (SD) | | 47.5% (11.5%) | 17.3% (10.2%) | 16.5% (8.8%) |
| **Regular Gait** | | | | |
| Total Steps | 31,401 | 29,993 | 26,155 | 28,433 |
| Mean Steps per Session | 1,047 | 1000 | 872 | 948 |
| % Mean Bias | | -4.5% | -16.8% | -8.5% |
| MAPE (SD) | | 6.4% (8.1%) | 16.3% (11.4%) | 9.2% (9.5%) |
| **Semiregular Gait** | | | | |
| Total Steps | 18,444 | 24,488 | 14,033 | 13,803 |
| Mean Steps per Session | 615 | 816 | 468 | 615 |
| % Mean Bias | | 32.8% | -23.9% | -25.2% |
| MAPE (SD) | | 33.5% (12.6%) | 23.9% (11.5%) | 25.8% (13.8%) |
| **Irregular Gait** | | | | |
| Total Steps | 4,542 | 25,404 | 4,625 | 3,121 |
| Mean Steps per Session | 151 | 847 | 154 | 104 |
| % Mean Bias | | 459.3% | +1.8% | -31.3% |
| MAPE (SD) | | 464.0% (84.3%) | 17.3% (14.1%) | 31.6% (15.9%) |

*MAPE (SD): Mean absolute percent error (standard deviation)*

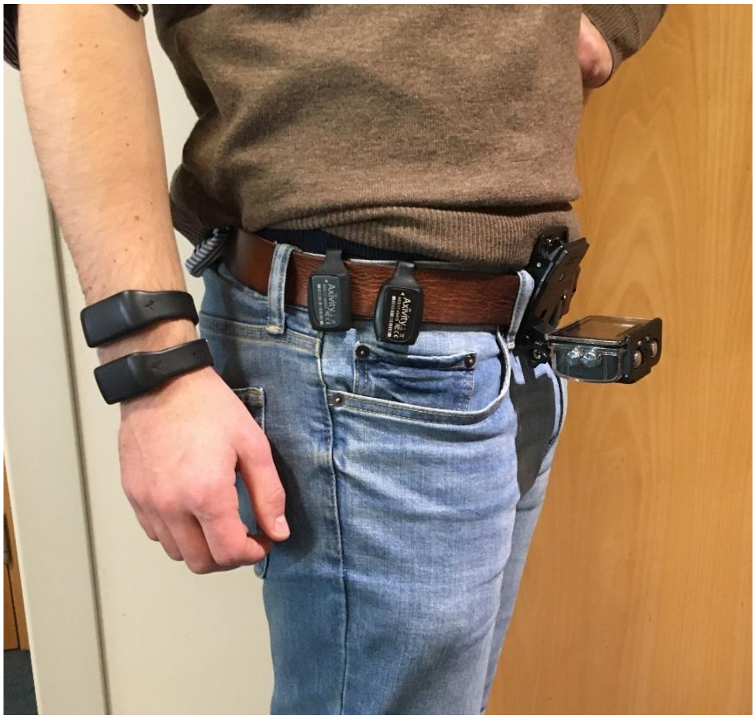

**Supplemental Figure 1: A foot-facing action camera clipped at the participant's waistline recording video for step count annotation. The participant is also wearing Axivity AX3 accelerometers at the dominant wrist and at the hip.**

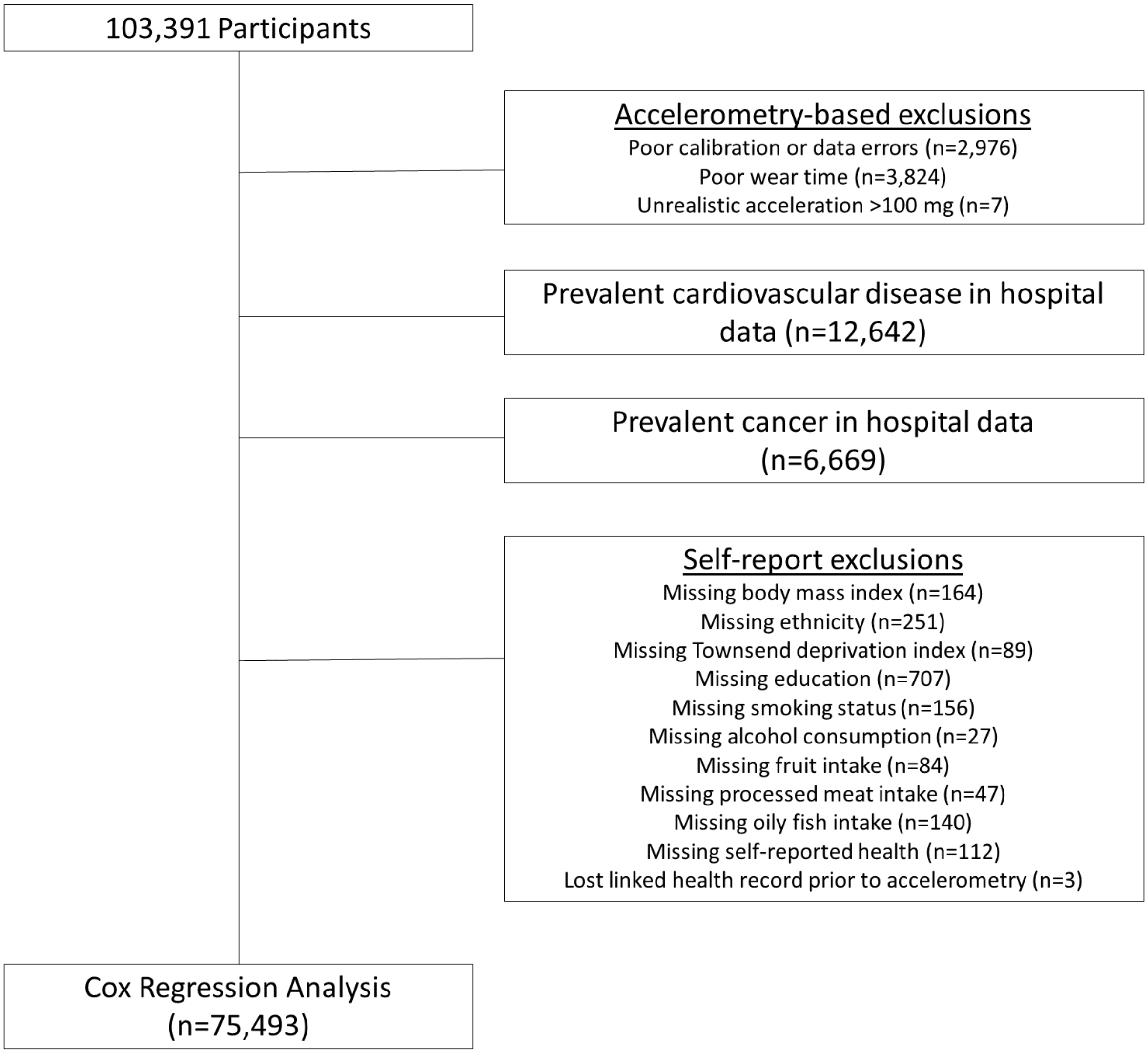

**Supplemental Figure 2: Flow diagram for descriptive and survival analyses in UK Biobank physical activity cohort.**

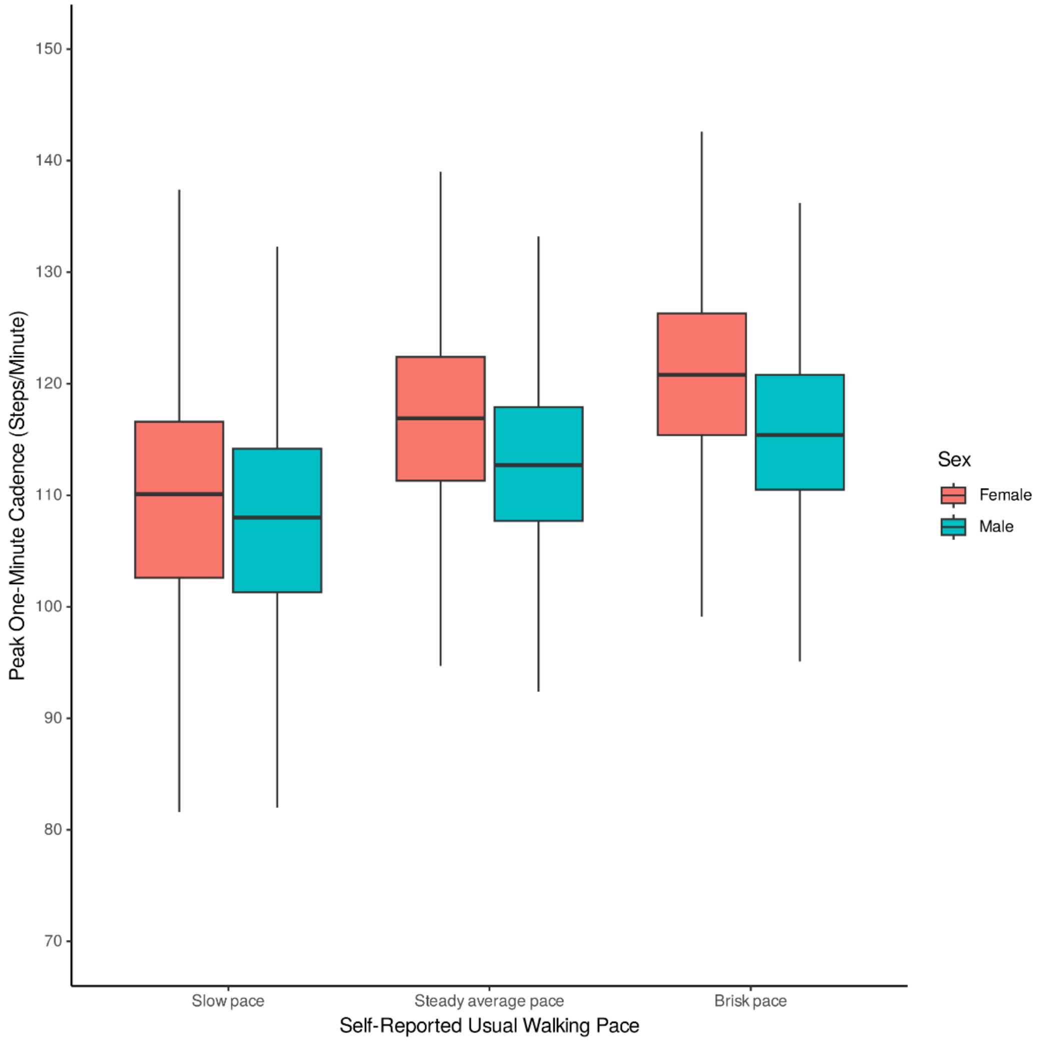

**Supplemental Figure 3: Peak step cadence in the UK Biobank Physical Activity Cohort, summarised by self-reported walking pace and sex.**

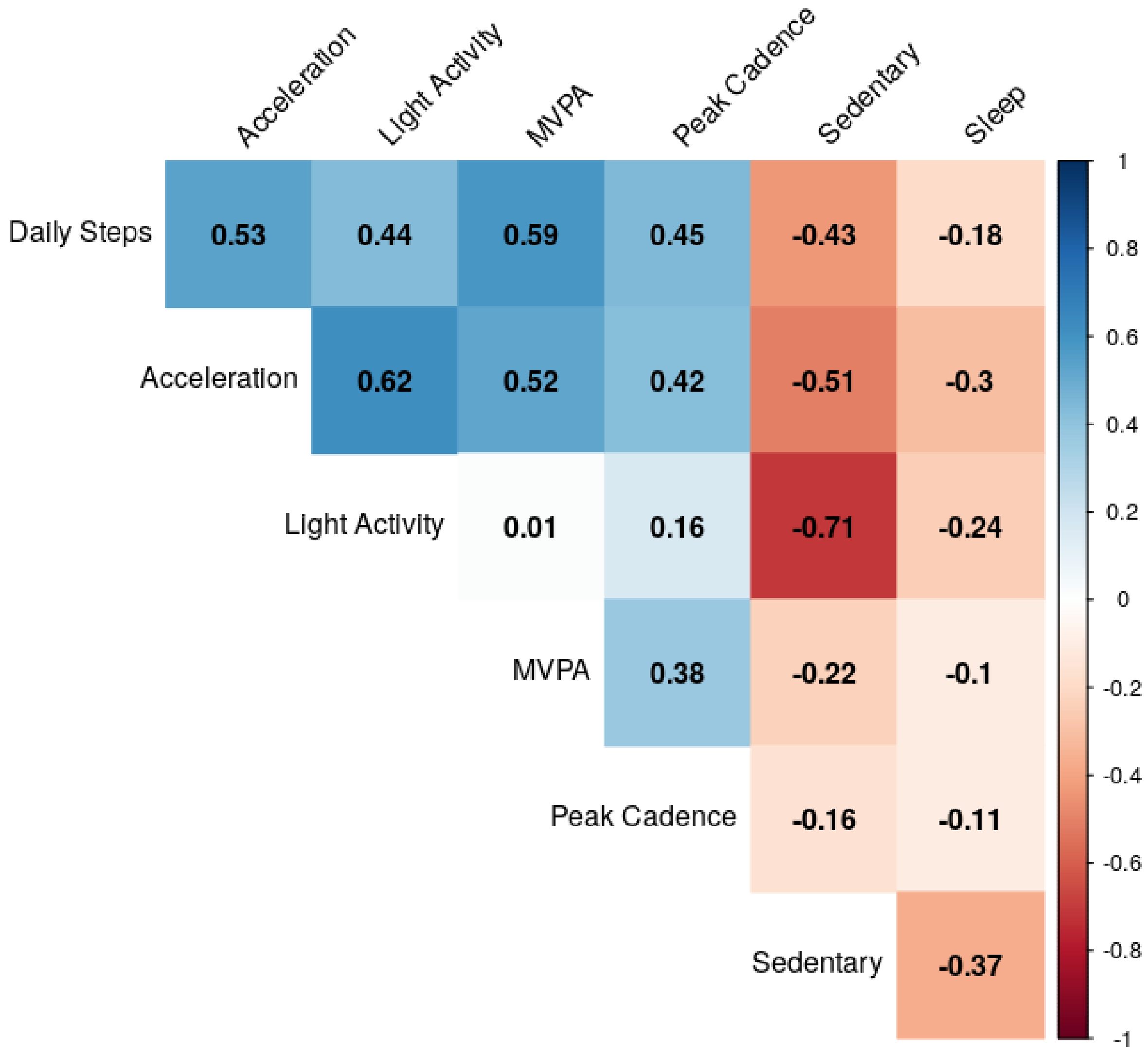

**Supplemental Figure 4: Spearman's rank correlation plot between accelerometer-based physical activity metrics, including overall acceleration, daily step count, and machine learning activity classifications of time spent in sleep, sedentary behaviour, light activity, and moderate-to-vigorous activity (MVPA) in 75,493 UK Biobank participants.**

**Supplemental Note 1: Comparative Step Detection Models**

Threshold-based step counting was conducted in R following code provided in Ducharme et al[8], wherein a single vector magnitude time series was generated for each participant with steps determined based on pre-defined acceleration thresholds following application of a $4^{th}$ order Butterworth 0.25-2.5 Hz band-pass filter[8]. The Verisense algorithm, an adapted version of a smartphone stepcounting algorithm by Gu et al.[34], was applied using the R script supplied via Github repository[25].

**Supplemental Note 2: Statistical Analysis, Further Detail**

To establish baseline activity levels within the UK Biobank, estimated marginal means of daily step count were calculated for participants when comparing activity relative to self-reported overall health status, adjusted for age and sex using the emmeans R package[35]. Estimated marginal means of daily step counts were similarly calculated for exemplar cohorts of participants with prevalent chronic renal failure, chronic obstructive pulmonary disease, depressive disorders, and insulin dependent diabetes, as defined by ICD codes listed in the Supplemental Materials. All survival models were implemented using v.3.2.11 of R's *survival* package[36]. Floating absolute risk for each step group was calculated v.2.47 of R's *Epi* package to allow a more clear assessment of the shape of association relative to the baseline step count quintile[37,38]. This method enables comparison of HR between any step count groups without specifying an arbitrary reference group. Smoothing spline plots were generated for the association of median daily step count with all-cause mortality and cardiovascular disease mortality from terms predicted using R's *termplot* function.

**Supplemental Note 3: Field and Code Usage**

**UK Biobank Fields**

UK Biobank Data Fields: 924 – Usual Walking Pace, 23101 – Body Mass Index, 21000 – Ethnic Background, 20116 – Smoking Status, 6138 – Qualifications, 1558 – Alcohol Intake Frequency, 189 – Townsend Deprivation Index at Recruitment, 2187 – Overall Health Rating, 31 – Sex, 52 – Month of Birth, 34 – Year of Birth, 90001 - Accelerometer Data cwa format (bulk), 90010 – Start time of wear (accelerometer), 90016 – Data quality, good calibration (accelerometer), 90015 – Data quality, good wear time (accelerometer) – Derived accelerometry: 40046, Sleep – Overall Average, 40047, Sedentary- Overall Average, 40048, Light – Overall Average; 40049 , Moderate-Vigorous – Overall Average

| **ICD9/10 Chronic Disease Codes (Hospital Episode Statistics)** | | |
|---|---|---|
| | **ICD9** | **ICD10** |
| Any Cardiovascular Disease or Associated Death | | I00-I78, I80-I99 |
| Any Cancer or Associated Death | | C00-C97 |
| Arterial Thromboembolism | 4442, 4448 | I740, I741, I742, I743, I744, I745, I748, I749 |
| Chronic Obstructive Pulmonary Disease | 4929 | J430, J431, J432, J438, J439, J440, J441, J448, J449 |
| Chronic Renal Failure | 5859 | N180, N181, N182, N183, N184, N185, N188, N189, N19 |
| Depressive Disorder | 3119 | F320, F321, F322, F323, F328, F329, F330, F331, F332, F333, F334, F338, F339 |
| Insulin-Dependent Diabetes | 25010, 25011, 25019 | E100-E109, E131, E141 |

| | ICD9 | ICD10 |
|---|---|---|
| Any Chronic Disease | 01192, 01199, 0130, 01619, 01729, 01789, 0703, 1800, 1809, 1830, 1860, 1869, 1413, 1416, 1419, 1420, 1440, 1505, 1519, 1521, 1530, 1532, 1533, 1534, 1536, 1537, 1539, 1540, 1541, 1542, 1543, 1551, 1561, 1570, 1574, 1579, 1590, 1600, 1610, 1613, 1619, 1623, 1629, 1649, 1702, 1707, 1709, 1720, 1723, 1725, 1726, 1727, 1729, 1730, 1731, 1732, 1733, 1734, 1735, 1736, 1737, 1950, 1951, 1740, 1743, 1744, 1745, 1748, 1749, 1799, 1859, 1890, 1882, 1889, 1906, 1909, 1913, 1916, 1919, 1939, 28099, 2820, 2829, 2830, 2849, 2860, 2864, 2869, 2870, 2871, 2872, 28730, 28739, 2875, 1359, 24499, 2410, 2411, 2419, 2420, 2422, 2423, 2429, 25010, 25011, 25019, 25000, 25001, 25009, 25029, 2503, 2504, 2505, 25099, 2520, 2530, 25329, 2550, 25549, 2722, 27249, 2910, 2918, 2953, 2959, 2961, 3119, 3000, 3027, 3015, 3320, 3321, 3335, 3336, 33379, 3338, 3339, 3409, 3418, 3419, 34510, 34519, 34550, 34559, 3459, 3460, 3461, 3462, 3468, 3469, 4359, 3479, 3550, 3551, 3555, 3556, 3558, 35809, 3593, 3594, 3421, 3429, 34300, 3431, 3439, 3441, 3446, 3449, 3314, 3360, 3369, 3352, 4019, 4039, 3969, 3940, 3942, 3949, 4240, 4241, 4139, 4109, 4129, 4280, 4281, 4151, 4442, 4448, 4538, 4254, 4273, 4270, 4271, 4272, 4274, 4276, 4278, 4279, 4141, 4373, | A150, B180, B200, C530, C56, C620, C01, C150, C160, C170, C180, C220, C23, C250, C260, C310, C340, C300, C400, C430, C440, C450, C460, C500, C540, C61, C64, C65, C670, C680, C690, C73, C810, B500, B550, D609, D630, D66, D690, D70, D860, E011, E040, E050, E100, E110, E140, E210, E220, E230, E240, E260, E271, E780, F000, F010, F020, F100, F110, F200, F21, F220, F250, F310, F320, F400, F420, F430, F450, F500, F520, F600, F640, G060, G10, G20, G300, G35, G360, G403, G430, G450, G470, G543, G600, G700, G710, G800, G910, G950, I10, I110, I081, I340, I350, I360, I371, I200, I210, I220, I252, I251, I500, I260, I740, I820, I630, I513, I610, I64, I420, I48, I440, I711, I050, J300, J310, J320, J370, J410, J430, J450, J47, J60, J848, J850, K210, K220, K221, K224, K225, K290, K580, K350, K400, K410, K420, K430, K440, K450, K500, K510, K552, K570, K760, K811, K860, K900, M0500, M1000, M1500, M5402, M750, M8008, N180, N200, N300, N310, N40, N411, N433, N600, N61, N701, N711, N72, N800, N810, N820, N920, N970, N46, N652, H800, H900, H250, H300, H400, H490, H530 |

| | |
|---|---|
| | 4410, 4411, 4414, 4416, 4423, 4429, 3989, 4778, 4779, 4720, 4721, 4722, 4730, 4731, 4732, 4733, 4739, 4760, 4910, 4912, 4918, 4929, 49309, 49319, 49390, 49399, 4949, 5019, 4952, 5159, 5109, 53010, 53011, 53019, 5300, 5305, 5302, 5310, 5311, 5314, 5315, 5317, 5319, 5320, 5321, 5323, 5324, 5325, 5326, 5327, 5329, 5334, 5339, 5343, 5344, 5349, 5350, 5351, 5353, 5354, 5355, 5356, 5368, 5369, 5400, 5401, 5409, 5419, 5429, 5500, 5501, 5509, 5520, 5530, 5521, 5531, 5522, 5532, 5523, 5533, 5528, 5538, 5539, 5550, 5551, 5552, 5559, 5620, 5621, 5771, 5790, 2749, 2320, 2321, 2322, 2323, 2324, 2325, 2326, 2327, 2329, 2330, 2331, 2332, 2333, 2334, 2335, 2336, 2337, 2339, 2340, 2341, 2342, 2343, 2344, 2345, 2346, 2349, 2350, 2351, 2352, 2353, 2354, 2355, 2356, 2359, 2361, 2362, 2363, 2366, 2369, 2380, 2381, 2382, 2383, 2384, 2385, 2386, 2387, 2389, 2390, 2391, 2392, 2393, 2394, 2396, 2399, 7170, 7171, 7260, 7261, 7262, 73305, 73309, 5859, 5920, 5921, 5929, 5950, 5951, 5952, 5953, 5958, 5959, 60099, 6011, 6039, 6100, 6101, 6102, 6103, 6104, 6108, 6109, 6110, 6141, 6151, 6160, 6170, 6171, 6172, 6173, 6176, 6178, 6179, 6180, 6181, 6184, 6186, 6187, 6188, 6189, 6190, 6191, 6192, 6198, 6199, 6261, 6262, 6264, 6265, 6266, 6267, 6268, 6269, 6280, 6282, 6283, 6284, 6288, |

| | |
|---|---|
| | 6289, 6069, 3811, 3812, 3813, 3821, 3822, 3823, 3831, 3850, 3851, 3852, 3853, 3860, 3861, 3863, 3869, 3872, 3879, 3890, 3891, 3892, 3899, 3662, 3665, 3668, 3669, 3632, 3638, 3650, 3651, 3652, 3655, 3656, 3659, 3780, 3781, 3784, 3785, 3788, 3789, 3682, 3684, 3688, 3689, 3690, 3696, 3697, 3699 |
